# Modeling aerial transmission of pathogens (including the SARS-CoV-2 virus) through aerosol emissions from e-cigarettes

**DOI:** 10.1101/2020.11.21.20235283

**Authors:** Roberto A Sussman, Eliana Golberstein, Riccardo Polosa

## Abstract

We examine the plausibility, scope and risks of aerial transmission of pathogens (including the SARS-CoV-2 virus) through respiratory droplets carried by exhaled e–cigarette aerosol (ECA). Given the lack of empiric evidence, we consider cigarette smoking and mouth breathing through a mouthpiece as convenient proxies to infer the respiratory mechanics and droplets sizes and their rate of emission that should result from vaping. To quantify direct exposure distance we model exhaled ECA flow as an intermittent turbulent jet evolving into an unstable puff, estimating for low intensity vaping (practiced by 80-90% of vapers) the emission of 6-200 (mean 79.82, standard deviation 74.66) respiratory submicron droplets per puff a horizontal distance spread of 1-2 meters, with intense vaping possibly emitting up to 1000 droplets per puff in the submicron range a distance spread over 2 meters. Since exhaled ECA acts effectively as a visual tracer of its expiratory flow, bystanders become instinctively aware that possible direct contagion might occur only in the direction and scope of the jet.

## 1. Introduction

There is currently a broad consensus, endorsed by the WHO Organization et al. (2020) and the CDC Brief (2020), that available data supports the occurrence of direct contagion of the SARS-CoV-2 virus by close range exposure to relatively large droplets emitted by infectious invididuals. While there is also a broad consensus on the factual occurrence of contagion through indirect exposure to smaller submicron droplets denoted by the term “aerosols” (see for example Liu et al. (2020); Li et al. (2020); Lu et al. (2020); Cai et al. (2020); Shen et al. (2020)), its scope and relevance still remains controversial Klompas et al. (2020); Morawska and Milton (2020); Morawska and Cao (2020); NAS (2020); Jayaweera et al. (2020); Shiu et al. (2019) ^1^ .

The evolution of bioaerosols spreading disease contagion through respiratory droplets has been widely studied, as can be appreciated in reviews on generic pathogens by Gralton et al. (2011); Zhang et al. (2015), the influenza and SARS viruses (Weber and Stilianakis (2008) and Yu et al. (2004)) and in spread risk modeling Sze To et al. (2008) (see the chapter on bioaerosols and cited references therein in Ruzer and Harley (2012)). As expected, the current COVID-19 pandemic has motivated the study of direct and indirect aerial transmission of the SARS–CoV–2 virus through various expiratory activities.

The purpose of the present paper is to fill an important gap in the above mentioned body of literature on pathogen (including SARS-CoV-2) spread through respiratory droplets, namely: to examine the plausibility, scope and risks of this transmission through a different expiratory route: exhaled e–cigarette aerosol (ECA). While there is no factual evidence nor proper elaborate research ^2^ on pathogen the transmission through this route, we hypothesize that its occurrance is entirely plausible simply because vaping (usage of e-cigarettes) is an expiratory activity (as well as smoking ^3^). To fulfill this task we develop in this paper a comprehensive and interdisciplinary theoretical modeling of the plausibility of this phenomenon. To compensate for the lack of empiric evidence we resort to cigarette smoking and mouth breathing through a mouthpiece as useful proxies to infer respiratory parameters and droplet emission that should occur through exhaled ECA.

The relevance of the present paper follows from the fact that the current COVID-19 pandemic has forced millions of vapers, smokers and non-users surrounding them into sharing indoor spaces under various degrees of home confinement. Objective research on COVID-19 transmission through exhaled ECA can serve to guide evidence based public policies to address public health concerns and risk management and minimization to address this phenomenon. Given the relevance of safety assessment in the context of the COVID-19 pandemic, we have elaborated a full risk analysis of vaping emissions in shared indoor spaces (Sussman et al. (2020)) and a short article (Sussman et al. (2021)) to discuss their public health and public policy implications.

It is necessary to issue the following important disclaimer: the present article is concerned only with the plausibility, scope and risks of pathogen (including SARS-CoV-2) transmission through exhaled ECA, not with vaping as a possible risk factor for becoming infected by the virus or for any evolution or stage of adverse health outcomes associated with COVID-19. Also, we will not be concerned with possible health hazards by users’ exposure to inhaled ECA or by-standers to exhaled ECA derived from the usage of e-cigarettes as substitute of tobacco smoking. Readers are advised to consult the available literature on these subjects (see extensive reviews Farsalinos and Polosa (2014); RCP (2016); McNeill et al. (2018); Daynard (2018); Polosa et al. (2019)).

In what follows we provide a section by section summary of the paper that illustrates its methodological structure. Background material is presented section 2: vaping styles and demographics (subsection 2.1); physical properties of inhaled and exhaled ECA (subsection 2.2); exhaled ECA as a marker of expiratory fluid flow (subsection 2.3). Section 3 (Methods) provides the material needed to infer respiratory mechanics of vaping (subsection 3.1), the effects of suction and mouthpieces (subsection 3.2) and ECA droplet emission from mouth breathing that should provide an appropriate proxy for vaping (subsection 3.3). In section 4 we present a model of a turbulent jet with finite injection evolving into an unstable puff to infer the horizontal distance that respiratory droplets carried exhaled ECA flow should be transported. The results obtained from these sections are summarized in section 5. Droplet emission (subsection 5.1): low intensity vaping should exhale 700 − 900 cm^3^ per puff, carrying 6–200 droplets (mean 79.92, SD 74.66), overwhelmingly in the submicron range, while high intensity vaping should exhale 1000 − 3000 cm^3^ per puff carrying possibly several hundreds and up to over 1000 droplets also in the submicron range. Distance for direct exposure: The jet/puff hydrodynamical model presented in section 4 yields a horizontal distance spread between 0.5 and 2 meters (low intensity vaping) and over 2 meters (high intensity) in the direction of the exhaled jet. We conclude the paper by presenting in section 6 its limitations, together with a final thorough discussion.

## 2. Background

### 2.1.. Vaping styles and demographics

#### 2.1.1. Puffing topography

Vaping is characterized by a wide range of distinct and individualized usage patterns loosely described by the parameters of *puffing topography*: puff and inter puff duration, puff volume and flow (see Dautzenberg and Bricard (2015); Farsalinos et al. (2018); Spindle et al. (2017); Soulet et al. (2019)). This style diversity complicates the study and evaluation of e–cigarette aerosol (ECA) emissions, more so given the need to upgrade standardization of vaping protocols, specially for the appropriate configuration of vaping machines used for research and regulation.

To simplify the description of vaping style, we consider two vaping topographies: (MTL), (DTL), described as follows

- Low intensity “Mouth–To–Lung” (MTL). It consists of three stages: (1) “puffing”, ECA is sucked orally while breathing through the nose, (2) the puffed ECA is withdrawn from the mouth held in the oropharyngeal cavity without significant exhalation and (3) inhalation into the lungs of the ECA bolus by tidal volume of air from mouth and nose inspiration. It is a low intensity regime involving low powered devices (mostly starting kits, closed systems and recent “pods”) roughly similar to the topography of cigarette smoking.
- High intensity “Direct–to–Lung” (DTL). As (1) in MTL but bypassing (2): the ECA bolus diluted in tidal volume is inhaled directly into the lung without mouth retention. It is mostly a high intensity regime associated with advanced tank systems.

The topography parameters characterizing these styles are listed in Table 1. It is important to remark that these parameters change when vaping *ad libitum* in natural environments instead of doing so in a laboratory setting. This was reported in Spindle et al. (2017): for example, average puff duration was about 20% longer ad libitum, 5 seconds vs 4 seconds in a laboratory setting.

**Table 1:** Parameters of vaping topography for vaping styles. Puff topography parameters: *m*_*b*_, *V*_*b*_, Φ_*b*_ are respectively mass (mg) (aerosol yield), volume (mL), flow (mL/sec) per puff of ECA bolus (aerosol yield). Notice that tidal volume *V*_*T*_ listed in the table is not the tidal volume for quiet rest breathing (400 − 600 mL), since vaping involves suction of ECA through a mouthpiece (see Sections 3.1 and 3.2). Puff time (secs) is *t*_*p*_. Values taken from from rough representative averages from data in figures 1 and 3 of Spindle et al. (2017) and also from Soulet et al. (2019).

| Parameters of vaping topographies. |  |  |  |  |  |
| --- | --- | --- | --- | --- | --- |
| Mouth to Lung (MTL) |  |  |  |  |  |
| Intensity | $m_b$ | $V_b$ | $\Phi_b$ | $t_p$ | $V_T$ |
| Low | 2–10 mg | 20–100 | 20–40 | 2–5 | 500–1500 |
| Direct to Lung (DTL) |  |  |  |  |  |
| Intensity | $m_b$ | $V_b$ | $\Phi_b$ | $t_p$ | $V_T$ |
| High | 10–40 mg | 300–500 | 100–300 | 3–6 | 1000–3000 |

A third puffing topography not included in Table 1 is “Mouth Puffing”: it shares step (1) of MTL but without step (3), with the ECA bolus diluted in tidal volume air being exhaled without lung inhalation. It is a low intensity regime but involving higher exhaled aerosol density, since less than 5% of aerosol mass is deposited in the mouth (see Asgharian et al. (2018)). Very few vapers and cigarette smokers use this style, but most smokers of prime cigars and tobacco pipes do.

#### 2.1.2. Demographics and markets

It is crucial to examine how representative among vapers are the different puff topographies and levels of intensity, something that has varied with time depending on the popularity and availability of different devices. Currently, low powered devices (mostly closed) are the most representative in the largest and most established markets. As shown in figure in Figure SM(1) (Supplemental Material with Credit to Eci (2020)) consumer surveys reveal that the overwhelming majority of vapers (90% in the USA and 80% in the UK) utilize low powered devices (mostly kits for beginners and closed systems) that operate with the low intensity MTL style, while only a minority use advanced open tank systems appropriate for the intense DTL style.

### 2.2. Inhaled and exhaled E-cigarette aerosol (ECA)

ECA is generated by various physicochemical processes: self–nucleated condensation in a super saturated medium initiates immediately once the e-liquid vapor leaves the coil, the nucleated centers generate small nm scale droplets that grow through coagulation and diffusion (see detailed explanation in Floyd et al. (2018)). The particulate phase is made of liquid droplets whose chemical composition closely matches that of the e-liquid: propylene glycol (PG), vegetable glycerin or glycerol (VG), nicotine, water Grégory et al. (2020), together with a trace level contribution of nanometer sized metal particles Mikheev et al. (2016). The gas phase is chemically similar. The aerosol contains nicotine and residues produced from the pyrolysis of the glycols and the flavorings (mainly carbonyls), which can be in either the gas or particulate phase depending on their vapor pressure and volatility Pankow (2017), with most of the PG evaporating into the gas phase and VG tending to be remain in the droplets Grégory et al. (2020).

Count mean diameter (CMD) distributions of mainstream ECA droplets vary depending on the device, puffing style of users, flavors and nicotine content Floyd et al. (2018); Lechasseur et al. (2019). Droplet number count is heavily dominated by submicron droplets with CMD distributions having either single modes below 100 nm or bimodal forms (one mode well below 100 nm and one in the range 100-300 nm) Floyd et al. (2018); Lechasseur et al. (2019); Scungio et al. (2018); Sosnowski and Odziomek (2018); Zhao et al. (2016); Fuoco et al. (2014). However, particle size grows with increasing coil power Lechasseur et al. (2019) and even in low powered devices the mass distribution is dominated by droplets larger than 600 nm Floyd et al. (2018). In fact, Floyd et al. (2018) found a third mode around 1 *µ*m that becomes more prominent at increasing power of the tested device while the nm sized modes decrease, likely because higher power involves larger vaporized mass that favors coagulation and scavenging of nm sized droplets by larger droplets.

The inhaled aerosol mass yield depends on the topography parameters given in Table 1. At inhalation of mainstream ECA instrument measured droplet density numbers are in the range *n* = 1–5×10^9^/cm^3^ (see Lechasseur et al. (2019); Scungio et al. (2018); Sosnowski and Odziomek (2018); Zhao et al. (2016); Fuoco et al. (2014)). Total average droplet numbers of *N*_*p*_ = 7.6×10^10^ were reported in Manigrasso et al. (2015) for a tank system using e–liquids with high nicotine content in a 2 second machine puff regime ^4^ with *V*_*b*_ = 50 mL puff volume (*N*_*p*_ decreases 25% with nicotine-free e-liquids). Using the same experimental design Fuoco et al. (2014) reported an increase of up to 30% for 4 second machine puff regime. The estimation *N*_*p*_ ∼ 10^10^ − 10^11^ is reasonable given a particle number concentration of ∼ 10^9^/cm^3^ and *V*_*b*_ = 20 − 100 mL of low intensity vaping, with *N*_*p*_ ∼ 10^12^ for high intensity vaping with *V*_*b*_ = 500 mL.

Data on the gas/particle phase partition of the aerosol mass yield *m*_*b*_ is roughly: 50% Total Particulate Matter (TPM), 40% PG/VG gas phase, 7% water vapor, < 3% nicotine McAdam et al. (2019), roughly a similar gas/particulate phase partition to that of tobacco smoke Martonen (1992). As shown in Grégory et al. (2020) and Pankow (2017) the presence of compounds in gas or PM form depends on their vapor pressure, with PG tending to be gaseous, VG in PM, for nicotine it depends on its PH, while some aldehydes (like formaldehyde) are most likely in the gas phase.

Values of particle numbers and densities for the exhaled ECA can be estimated by considering its retention by the respiratory system. Compound specific retention percentages were reported in St. Helen et al. (2016) for a wide variety of devices and e–liquids: 86% VG, 92% PG, 94% nicotine, while Samburova et al. (2018) reported 97% total aldehyde retention. This high retention percentages are consistent with the mass distribution of inhaled ECA dominated by larger micron sized droplets which tend to be efficiently deposited in the upper respiratory tracts Floyd et al. (2018). Assuming equal retention rate for the particulate and gas phases, we take as total mass of exhaled aerosol and total numbers of exhaled ECA droplet to be 10% of the values of *m*_*b*_ listed in Table 1 and 10% of the values of *N*_*p*_ = 6.7 × 10^10^ reported in Manigrasso et al. (2015) for a 2 second machine inhalation puff and 50 mL puff volume. Droplet number density of ECA as it is exhaled can be estimated from these values of *N*_*p*_ bearing in mind that the exhaled ECA is now diluted in tidal volumes *V*_*T*_ listed in Table 1 for the various vaping topographies. This yields number densities in the approximate range *n*_*p*_ = 10^6^ − 10^7^ cm^−3^ (lower to higher vaping intensities).

Exhaled ECA dilutes and disperses very fast. Its chemical composition is similar to that of inhaled ECA, both in the gas phase and the droplets Floyd et al. (2018), with PG and water in the latter evaporating rapidly. Since hyperfine nm sized droplets deposit efficiently by diffusion in the alveolar region and larger micron sized droplets (which tend to grow from hygroscopic coagulation Asgharian et al. (2018); Floyd et al. (2018)) deposit by impaction in the upper respiratory tracts Asgharian et al. (2018); Manigrasso et al. (2015); Lechasseur et al. (2019); Sosnowski and Odziomek (2018), the CMD distribution of ECA as it is exhaled should be dominated by modes in intermediate ranges 0.1 − 0.5 *µ*m. Since there are no ECA measurements at the exhalation point (the vaper’s mouth), we can estimate the representative droplet diameter by a rough order of magnitude calculation: assuming an aerosol mass yield of 5 mg of inhaled ECA for a low powered device, a 90% retention of aerosol mass with 50% made of PM, the total droplet mass of exhaled ECA should be around *M*_*p*_ = 0.25 mg. Assuming a 90% retention of inhaled droplets, the total number of exhaled droplets should be *N*_*p*_ = 7.6 × 10^9^ droplets (from inhaled numbers in Manigrasso et al. (2015)), leading to a median droplet mass of 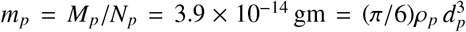, where *ρ_p_* is the droplets density that we can assume to be close to VG density: ρ_*p*_ = 1.3 gm/cm^3^, leading to *d*_*p*_ = 0.38 *µ*m. Similar order of magnitud values are obtained for the parameters of high intensity vaping.

The fact that CMD chamber measurements are in the range *d*_*p*_ = 0.1−0.2 *µ*m can be explained by the fact that detectors are located 1-2 meters from the exhalation source, thus measured ECA droplets have already undergone significant degree of dilution and evaporation (as shown in Grégory et al. (2020) droplets’ mass can decrease by one third in just 1 second by evaporation of its PG content). This is consistent with droplet number densities dropping at least two orders of magnitud from ∼ 10^6^ − 10^7^ cm^−3^ as they are exhaled to *n* ∼ 10^4^ − 10^5^ cm^−3^ at one meter distance from the emission and further dropping to near background levels *n* ∼ 10^3^ cm^−3^ at two meters Zhao et al. (2017); Martuzevicius et al. (2019); Palmisani et al. (2019).

### 2.3. Exhaled ECA as a visual tracer of respiratory fluid flow

The overwhelmingly submicron droplets that form the particulate phase of ECA have negligible influence on its fluid dynamics, acting essentially as visible tracers or (to a good approximation) as molecular contaminants carried by the fluid. This follows from its basic fluid dynamical characteristic: it is a “single–phase fluid flow” (SFF) system Yeoh and Tu (2019); Elghobashi (1994). As a consequence, ECA droplets visually mark the actual expiratory flow associated with vaping exhalations (we discuss the optical properties that allow for its visualization in Sussman et al. (2020)).

Exhaled ECA is just one among numerous gas markers and aerosols in a SFF regime that serve (and are widely used) to visualize expired air Ai et al. (2020); Nazaroff (2004). This also applies to mainstream exhaled tobacco smoke, whose particulate matter is also made of submicron liquid and solid droplets. In fact, there are studies that have directly used cigarette smoke as a tracer to visualize respiratory airflows Gupta et al. (2009, 2010); Ivanov (2019). Respiratory droplets potentially carried by exhaled ECA would not change its possible role as a tracer of expiratory flows, since as we show further ahead (section 3.3) these droplets are also overwhelmingly in the submicron range and their numbers are several orders of magintude fewer than ECA droplets.

Submicron ECA droplets are carried by a fluid made of the gas phase of ECA strongly diluted in exhaled air (in practice, we can think of the carrier fluid as exhaled air at mouth temperature ∼ 30 − 35° C and 80-100% relative humidity). Under such conditions ECA droplets essentially follow the fluid flow because of their little inertia, as they are well within the Stokes regime with Reynolds numbers *Re*_*p*_ ≪ 1 and negligibly small relaxation times *t*_rel_, the response time of an aerosol particle to adjust to external forces. For *d*_*p*_ = 0.3 *µ*m we get Hinds (1999)

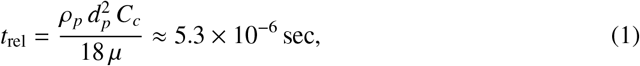

where *ρ_p_* ∼ 1.3 gm/cm^3^ (close to VG density), *µ* = 1.895 × 10^−5^gm/(sec cm) the dynamic viscosity of air at 35 C and *C*_*c*_ = 1+(λ/*d*_*p*_)[2.34+1.05 exp(–0.39*d*_*p*_/λ)] ≈ 1.4 is the Cunningham slip factor with *λ* = 0.066 *µ*m the mean molecular free path of air.

The relaxation time (1) provides the time scale for a particle released into a fluid with velocity *U* along a horizontal stream to settle into the fluid velocity (neglecting gravity). In this case (see Chapter 3 of Hinds (1999)) the velocity of the particle 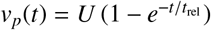 becomes practically identical to *U* in about 10^−5^ seconds (instantaneously in practical terms), thus justifying the notion of particles simply following the fluid flow with (practically) no influence on its dynamics.

This behavior occurs also for the larger ECA droplets of *d*_*p*_ ∼ 1 *µ*m whose relaxation times are *t*_rel_ ∼ 10^−4^ (since 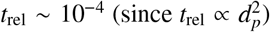). Evidently, these relaxation times are much smaller than macroscopic characteristic times of the carrier fluid (for example a 2 second inhalation time or even the tenths of a second the ECA stays in the mouth cavity Asgharian et al. (2018)). The Stokes number is defined as *S t* = *t*_rel_/*t*_*f*_, where *t*_*f*_ is a characteristic fluid time, hence for the exhaled ECA we have *S t* ≪ 1, which is another criterion to define SFF systems.

Evidently, larger droplets (diameters larger than a few *µ*m) are present in particle diameter distributions (of both ECA and respiratory droplets) and such particles should contain a significant portion of the aerosol mass Floyd et al. (2018), but they are too few in numbers and deviate from the flow following ballistic trajectories, thus do not affect the dynamics of the carrier fluid to consider ECA as a biphasic fluid flow system (as is the case with violent coughs or sneezes, see Bourouiba et al. (2014)).

## 3. Methods: Inferences on respiratory droplets spread by ECA

### 3.1. Vaping as a respiratory process

Tobacco smoke is a valid physical reference for ECA, as it is also an aerosol in a SFF regime characterized by a particulate phase made of predominantly submicron particles with similar particle numbers and diameter distributions Floyd et al. (2018); Sosnowski and Odziomek (2018); Sosnowski and Kramek-Romanowska (2016) (though the particulate and the gas phases of each aerosol have very different chemical properties). This fact, together with the fact that most vapers are either cigarette smokers or ex–smokers of cigarettes, justifies inferring the respiratory parameters of vaping (especially exhaled volume) from the respiratory parameters of smoking reported in the literature (see reviews in Bernstein (2004); Marian et al. (2009), see also a summary of studies in Table SM(2), Supplemental Material SM(2)).

#### 3.1.1. Respiratory parameters of smoking

While there is a wide individual diversity in respiratory parameters among smokers, roughly the same three puffing topography patterns identified in section 2.1 for vaping occur in smoking (with tobacco smoke instead of ECA) Higenbottam et al. (1980). As with vaping, the most common cigarette smoking topography is MTL, an expected outcome since most vapers are either ex-smokers or current smokers of cigarettes. While a sizable minority of 10-20% of vapers (see Section 2.1.2) follow the DLT style, the vast majority of smokers avoid direct lung inhalation because it is too irritant Tobin et al. (1982b) (and is consistently associated with airways narrowing Higenbottam et al. (1980)). In fact, avoidance of the direct lung inhalation of DTL style is very likely an organic response to minimize to a tolerable level the irritant quality of tobacco smoke Higenbottam et al. (1980); Tobin et al. (1982b,a).

Regarding its respiratory parameters, cigarette smoking involves a larger percentage of vital capacity than rest breathing: 20-25% Bernstein (2004), though low intensity inhalators might use on average only 14% Tobin et al. (1982a). It is extremely likely that these figures apply at least to MTL vaping. Other parameters such as expired tidal volume, puff times and volumes obtained in observational studies are listed in Table **??**, where we used outcomes from references cited in two comprehensive reviews Bernstein (2004); Marian et al. (2009). The summary of these outcomes is roughly (see list of studies in Table SM(2), Supplemental material):

- Puff Volume” (volume of the smoke bolus drawn from the cigarette) 20-70 mL,
- Puffing Times (time to draw the smoke bolus from the cigarette) ∼ 2 seconds
- Total smoking time lapses (inhalation, breath hold and exhalation) ∼ 4 − 7 seconds
- Tidal volumes (the volume of the total inhaled/exhaled smoke mixed with air, *V*_*T*_ in table 1) vary widely between 500 and 1500 mL (with some outliers reaching as low as 300 mL and as high as 2000 mL), but typically group averages are between 700 and 900 mL.

It is worth remarking that puffing times are slightly shorter but roughly comparable to those of MTL vapers, while tidal volumes are 25-30% larger than rest tidal volumes (400-600 mL), though the measurement of these volumes is subject to at least a 10% error Herning et al. (1983) and also, not all air drawn with the purpose of inhaling smoke is actually inhaled. Most studies report inhaled volumes, but exhalation volumes are roughly comparable (see Table **??**), as smoke is highly diluted in air and its retention barely affects volume measurement.

#### 3.1.2. Suction

As opposed to rest breathing, smoking and vaping involve suction: the inward force needed to draw smoke (or ECA) associated with the negative/positive pressure gradient Δ*P* generated by the diaphragm driven expansion/contraction of the lungs. Airflow resistance follows from the relation between the flow of air volume 𝒬 = *dV*/*dt* and this pressure gradient, a relation that can be modeled by the following power law (see Wheatley et al. (1991); Jaeger and Matthys (1968))

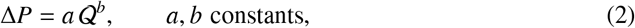

where *a, b* are determined empirically. This power law can be related to fluid dynamics (see discussion in Jaeger and Matthys (1968)): the constants *a* and *b* correlate with fluid density, while the exponents *b* can be referred to the “classical” flow regimes: *b* = 1 corresponds to laminar flow with Reynolds numbers *Re* < 10 (Pouseuille law), *b* = 1.75 to turbulent flow *Re* ∼ 10000 (Blasius law) and *b* = 2 is the “orifice” flow characterized by turbulent flow in narrow pipes and containers.

The theoretical connection with fluid mechanics has motivated airflow resistance measurements in the upper respiratory system that yield values around *b* = 1.84 Wheatley et al. (1991); Jaeger and Matthys (1968) for resting oral and nasal breathing. An excellent fit of this power law relation to the classical orifice flow *b* = 2 was found for a conventional cigarette and a two second generation e–cigarettes by Sosnowski and Kramek-Romanowska (2016), with the e–cigarettes flow resistance *a* between 3-4 times larger than the conventional cigarette. As a consequence, given the same suction effort (same Δ*P*) a conventional cigarette yields a puffing flow 𝒬 between 3-4 times larger than the tested e–cigarettes (second generation). However, vapers can compensate the higher flow resistance of ECA and draw relatively large aerosol mass with the same suction effort by puffing for longer times (as shown by topography studies). Also, the laboratory measurements of Sosnowski and Kramek-Romanowska (2016) were conducted under idealized conditions and are very likely to vary among the many e-cigarette devices in natural usage conditions.

A factor that distinguishes cigarette smoking from vaping is that the latter involves suction of ECA through a mouthpiece. However, in most of the studies listed in Table SM(2) (Supplemental Material) the subjects smoked through cigarette holders that are part of the laboratory instrumentation. This makes the listed outcomes more useful to infer respiratory parameters for vapers, at least for those vaping in the MTL style, since these holders are of similar size and shape as the narrow e–cigarette mouthpieces. Though, usage of cigarette holders does not seem to introduce significant changes in tidal volume, as can be seen by comparing outcomes from studies that used holders with those who did not in Table SM(2) (we comment further on the effect of mouthpieces in Section 3.2).

Since MTL is the most common topography among smokers and vapers (most of whom are ex-smokers or current smokers), we can assume that MTL style vaping is characterized by qualitatively similar puffing and respiratory parameters to those listed in Table SM(2). While some smokers inhale without a mouth hold as in DTL style, this does not seem to involve in them a significantly higher tidal volume, most likely because it can be too irritant Higenbottam et al. (1980); Tobin et al. (1982a). The lesser irritant nature of ECA is a plausible explanation for a larger proportion of vapers that can tolerate DTL topography, which means suction of a much larger aerosol mass Soulet et al. (2019); Cahours and Prasad (2018) and thus significantly larger puffing and tidal volumes than in MTL style (made easier by usage of high powered devices). A puff volume of 500 mL can yield under idealized laboratory conditions an inhalation tidal volume close to 3 LT Vas et al. (2015), which justifies the more plausible values listed in Table 1.

### 3.2. Mouthpieces, noseclips and the breathing route

Mouthpieces (MP) and nose-clips (NC) (to block nasal inspiration) are standard instruments in observational studies, not only those aimed at studying droplet emission, but of respiratory patterns and flows in human subjects. Since the results of these studies can serve as appropriate proxy values to infer droplet emission in vaping, it is important to assess the effects of these instruments in respiratory mechanics. For the purpose of the present article, this issue is interesting because ECA is inhaled in e–cigarettes through mouthpieces (though without obstruction of nasal breathing).

#### 3.2.1. Observational data on breathing through mouthpieces and noseclips

Several studies conducted in the 1970’s and 1980’s (Gilbert et al. (1972); Askanazi et al. (1980); Hirsch and Bishop (1982); Douglas et al. (1983); Weissman et al. (1984)) have shown that breathing through MP’s and NC’ affect all respiratory parameters with respect to unencumbered nose breathing: while tidal volume increases roughly 20% with respect to its normal rest value of 400-600 mL in all studies, inhalation and exhalation times and respiratory frequency are much less affected. It was shown by Weissman et al. (1984) that a NC without a MP produces a similar increase of tidal volume but also significant increase of inhalation times (15%) and exhalation times (22%). In two of the studies (Askanazi et al. (1980); Weissman et al. (1984)) the subjects were in supine position, which yields slightly smaller tidal volumes but does not modify inspiration/expiration times (Kera and Maruyama (2005)).

Besides possible reasons like the psychological sensorial stimulation of receptors by colder air in mouth inspiration and the stress of breathing through instruments, another possible explanation for the observed change in respiratory parameters of MP’s is the change of airflow resistance, for example: a 70–90% reduction seen by Weissman et al. (1984) brought by the large added mouthpiece dead space (up to 80 mL), while the larger airflow resistance from the standard 17 mm to a narrower 9 mm MP (closer in size to mouthpieces used in vaping) reduced the increase of tidal volume to 11% and inhalation/exhalation times to 9% Weissman et al. (1984). Therefore, the MP’s of e-cigarettes should produce similar modifications of respiratory parameters as with the narrower MP.

#### 3.2.2. Effects of the breathing route

In the studies discussed above there was no separation between usage of instruments (MP & NC) and oral breathing. Rodenstein, Mercenier and Stanescu Rodenstein et al. (1985) conducted several experiments with 14 healthy subjects with the aim of looking separately at the effects of MP’s and a NC’s. Their main result is that changes of respiratory parameters (rough 20% and 10% increase of tidal volume and inhalation/exhalation cycle) are entirely due to the forced oral breathing induced by the NC, in fact, nose occlusion is not even necessary to produce these changes: it is sufficient to simply instruct the subjects to breath through the mouth to observe an increase the tidal volume by a similar proportion as with the use of a NC: from 456 ± 142 to 571 ± 199 mL, though inhalation/exhalation times and other parameters remain almost the same (likely because of breathing without instrumentation).

The physiology behind the effects of the breathing route is similar to the one discussed in the study of pipe and cigarette smokers Rodenstein and Stănescu (1985): changes of respiratory parameters depend on the degree with which subjects are able to maintain air flowing through the nose. These parameters exhibit minor variation as long as this air flow is not occluded and the oropharyngeal isthmus remains closed. The parameters change significantly when nose occlusion separates the soft palate and the tongue and opens the oropharyngeal isthmus to allow air to flow entirely through the mouth. However, after the initial puffing, air flows through both nose and mouth in smoking and vaping (except the Mouth Puffing style), with the soft palate closing and rising enough to control the oral or nasal flow. While vaping involves usage of a MP, it does not involve nasal occlusion, hence the increase of expired tidal volume with respect to rest breathing (and puffing times respect to smoking) should be primarily due to suction of ECA.

### 3.3. Likely characteristics of respiratory droplets carried by ECA

The discussion in the previous sections has allowed us to infer the characteristics and parameters of the respiratory mechanics of vaping. We need now to identify among respiratory processes the ones that most closely fit these parameters in order to use their available experimental data to infer the capacity of vaping for respiratory droplets emission.

#### 3.3.1. The right respiratory proxy: mouth breathing

Intuitively, vaping as a respiratory process should be close to mouth breathing, as both involve a roughly time-symmetric cicle of inspiration/expiration (as opposed to vocalizing, coughing or sneezing). Given the fact that exhaled ECA is a single phase flow (SFF) system (see section 2.3), another good criterion to relate vaping to mouth breathing is the comparison between their fluid exhalation velocity *U*_0_ and measured analogous velocities in other respiratory processes.

The exhalation velocity *U*_0_ can be roughly inferred qualitatively by considering an exhaled tidal volume of fluid flowing through the respiratory tracts. Considering the respiratory parameters discussed in the previous sections (summarized in Table 1) we can use the simple approximate formula

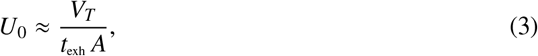

where *V*_*T*_ is the expiration tidal volume (in cm^3^), *t*_exh_ is the exhalation time in seconds and *A* is the combined mouth and nose area (in cm^2^), as the fluid carrier of both ECA and tobacco smoke is exhaled through the mouth and nose. From the values listed in Tables 1 and **??** we have:

- MTL vaping and smoking: *V*_*T*_ = 500 − 1500 mL and *t*_exh_ = 2 − 3 sec., while values for the combined mouth/nose area has been measured between *A* = 2 − 3 cm^2^ Gupta et al. (2010).
- DTL Vaping: *V*_*T*_ = 1500 − 3000 mL with *t*_exh_ ≈ 3 − 4 sec. and *A* ≈ 3 cm^2^. Given the large amount of exhaled fluid we assume longer exhalation times and larger mouth opening area.

From the combination of the parameter values mentioned above we have

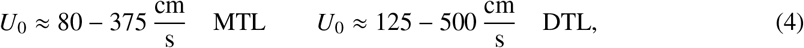

which reinforces the intuitive notion that mouth breathing is the appropriate respiratory proxy for MTL and DTL vaping (and also cigarette smoking), since these estimated exhalation velocities are well within the range of those of exhaled breath in mouth breathing without nose occlusion by NC’s Xu (2018); Xu et al. (2015, 2017), which have been estimated and measured by various techniques (including Schlieren photography). Exhalation velocities in the most intense DTL vaping regime approach in their upper end the velocities of vocalizing but fall short of those of coughing and sneezing. As a reference, measurements of *U*_0_ using Particle Image Velocimetry resulted in averages of 3.9 m/s for speaking and 11.7 m/s for coughing Chao et al. (2009) (measurements in Zhu et al. (2006) resulted in 6-22 m/s with average 11.2 m/s for coughing), while 35 m/s has been estimated for sneezing Chen and Zhao (2010); Wei and Li (2016); Scharfman et al. (2016).

#### 3.3.2. Data on droplet emission from mouth breathing

There is an extensive literature on respiratory droplets emitted by mouth breathing at different levels of lung capacity, including rest tidal volume breathing (< 20% of vital capacity). We list a selection of these studies in Table 2, as they are the ones that can serve as proxies for vaping and smoking (at least MTL style). In practically all the listed studies subjects breathed through MP’s (mouthpieces) and NC’s (noseclips), which as discussed in section 3.2, involves occlusion of nasal air flow that implies a slightly modified mechanics and about 20% larger tidal volume with respect to normal unencumbered breathing.

**Table 2:**
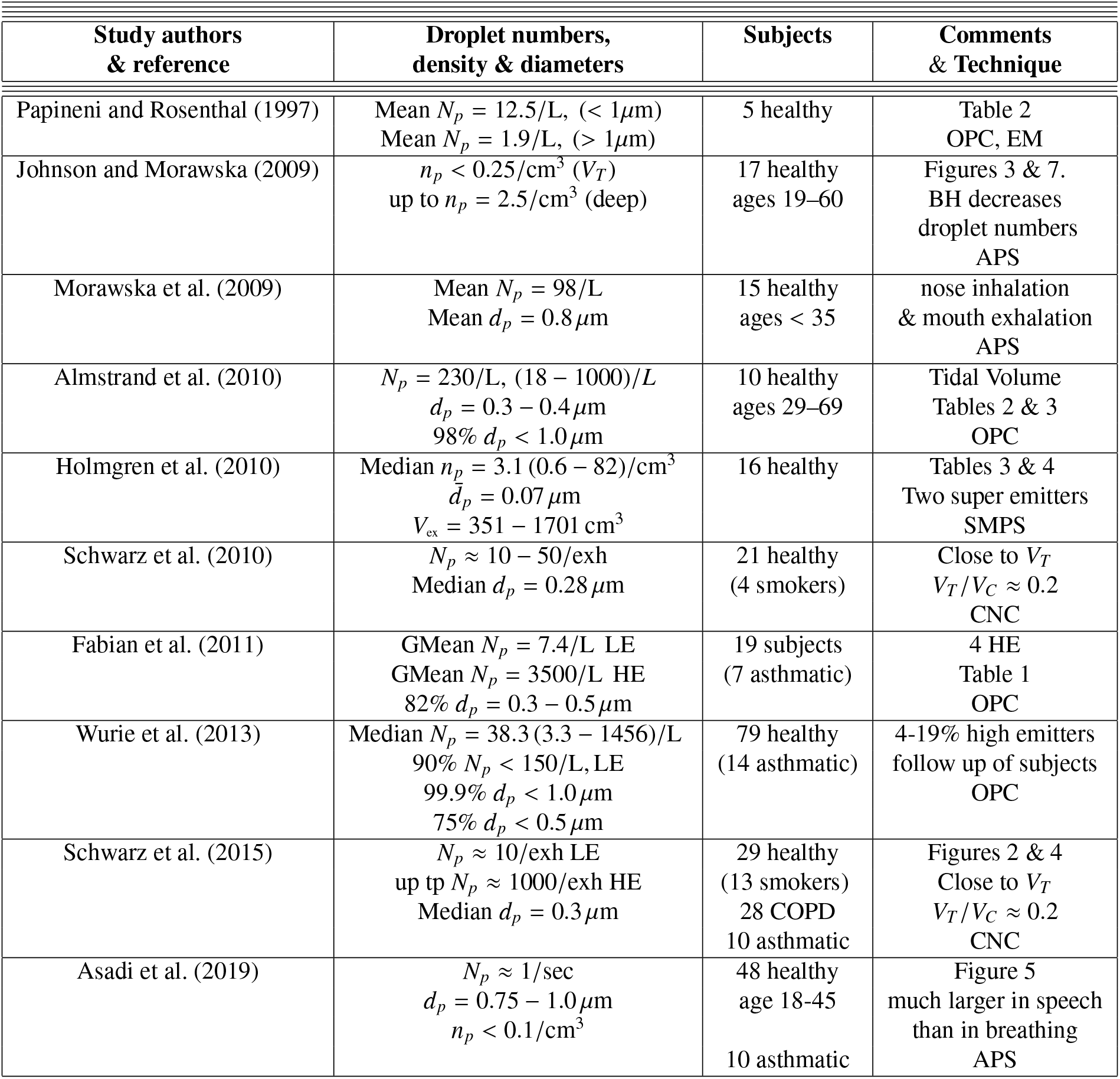
Droplet emissions for mouth breathing and tidal volume. The symbols *N*_*p*_ and *n*_*p*_ stand for droplet number per exhalation and average droplet number density (cm^−3^). LE, HE, BH, L, and exh are Low emitters, High Emitters, Breath Hold, litter and exhalation. The acronyms OPC, EM, APS, SMPS, CNC stand for Optical Particle Counter, Electron Microscopy, Aerodynamic Particle Sizer, Scanning Mobility Particle Sizer, Condensation Nucleus Counter. The subjects in all studies (save possibly Johnson and Morawska (2009)) breathed through a mouthpiece wearing a noseclip.

The fact that emitted respiratory droplets in tidal volumes close to rest breathing are overwhelmingly in the submicron range (as shown by Table 2) implies a very rapid evaporation (0.01 sec) that in practice can be considered as instantaneous, with the emitted droplets being overwhelmingly desiccated droplet nuclei made of salt crystals and lypoproteins and being about roughly half their original diameter (see Nicas et al. (2005)). The exhaled breath will also contain some larger particles *d*_*p*_ ∼ 1 − 3 *µ*m that evaporate in timescales of 0.1 sec so that (given the exhalation velocities in (4)) they become desiccated nuclei at horizontal distances of 10-30 cm. As a consequence, relative humidity bears negligible influence on the evolution of the bulk of emitted droplets.

While some of the studies in Table 2 were motivated by investigating droplet emission in the context of airborne pathogen contagion (for example Papineni and Rosenthal (1997); Fabian et al. (2011); Wurie et al. (2013); Asadi et al. (2019)), the motivation of others is to probe various mechanisms of droplet formation (Johnson and Morawska (2009); Morawska et al. (2009); Almstrand et al. (2010); Holmgren et al. (2010); Schwarz et al. (2010, 2015), see comprehensive discussion and reviews in Wei and Li (2016); Bake et al. (2019); Haslbeck et al. (2010)), specifically the airway reopening hypothesis of small peripheral airways that normally close following a deep expiration, which was further tested by computerized modeling by Haslbeck et al. (2010) who simulated this mechanism of particle formation by rupture of surfactant films involving surface tension. The mechanism was probed by Johnson and Morawska (2009) by showing that concentrations of exhaled particles significantly increase with breathing intensities higher than rest tidal volume, but also for fast exhalations but not fast inhalation, while droplet numbers increased up to two orders of magnitude: from ∼ 230/LT in tidal volume (0.7 Lt) to over 1200/LT in a breathing maneuver from fractional residual capacity to total lung capacity (see Almstrand et al. (2010)).

The difference in droplet formation between breathing and speaking was examined by Johnson et al. (2011): normal and deep tidal breathing produced submicron distributions related to those of other studies probing the airway reopening mechanism, while speech and cough produced larger diameter modes (∼ 1 *µ*m) with particle formation associated with vocal cord vibrations and aerosolization in the laryngeal region. A third mode of median diameters of 200*µ*m was associated with the presence of saliva between the epiglottis and the lips.

Breath holding between inspiration and expiration were found by Johnson and Morawska (2009) to significantly reduce concentrations of exhaled droplets in proportion to the breath hold time. The same outcome was found by Holmgren et al. (2013) for inspiration to total lung capacity, but droplet numbers increased when the breath hold occurs before inspiration. These outcomes fit predicted effects of gravitational settling in the alveolar region. Since the observations of Johnson and Morawska (2009); Holmgren et al. (2013) involved breathing intensity well above tidal volume up to total vital capacity, it is not possible to compare them quantitatively with the breath hold of the MTL style. However, gravitational settling of larger droplets must also occur in the bucal cavity under normal vaping conditions (see Asgharian et al. (2018)), so it is reasonable to assume that reduction of exhaled droplet numbers should also occur at lower intensity in MTL style vaping.

## 4. Hydrodynamical modeling of direct exposure

In the previous sections we have inferred the submicron characteristics and rate of emission of respiratory droplets expected to be carried by exhaled ECA. We need to estimate now how far can these respiratory droplets be carried to evaluate the distance for direct exposure of bystanders to pathogens potentially carried by these droplets

Exhaled ECA is injected into surrounding air a given horizontal distance roughly in the direction of the exhaled flow. Since it involes a finite fluid mass of a SFF aerosol during a finite injection time (exhalation time), the appropriate dynamical model for it is a turbulent puff with a starting momentum dominated jet that lasts while the fluid injection is on (see Pope (2001); Rajaratnam (1976); Abani and Reitz (2007); Abraham (1996); Ghaem-Maghami (2006); Ghaem-Maghami and Johari (2010); Sangras et al. (2002, 2003)). A schematic description of this system is furnished by Figure 1. We will not be concerned with the few larger particles (diameters *d* ∼ 1 − 5*µ*m and over) that initially follow the fluid stream but (depending on their size) exit the main flow to follow ballistic trajectories until they either deposit on surfaces, settle on the ground or evaporate (see Wei and Li (2016); Bourouiba et al. (2014)).

**Figure 1:**
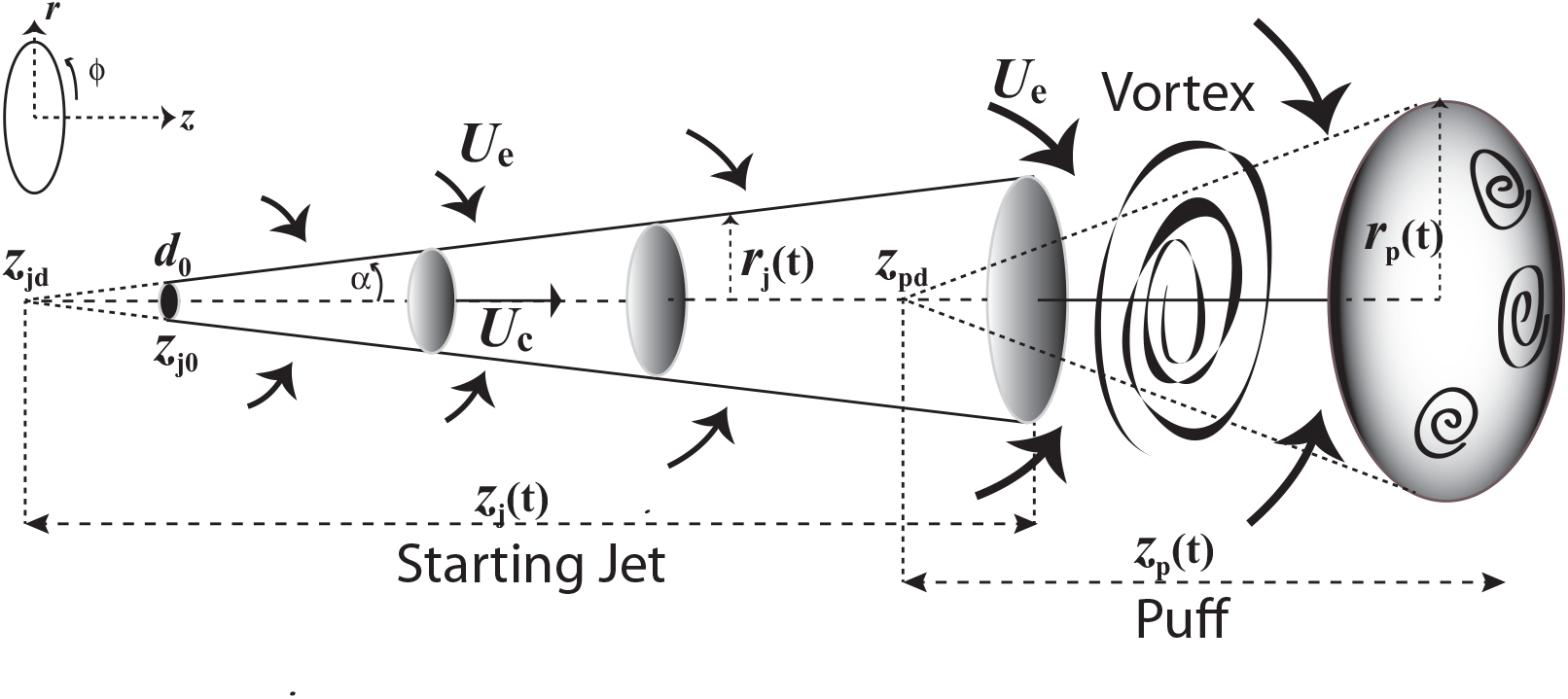
Puff and initial Jet with axial symmetry. The stating jet is propelled by linear momentum parallel to the centerline velocity *U*_*c*_, the arrows above and below represent the entrainment velocity mixing surrounding air with the carrier fluid. As the fluid injection terminates (end of exhalation), the entrained air makes about 40% of the fluid mass making the the transition into a ellipsoidal puff through highly turbulent vortex structures. At this point the puff is likely to disperse rapidly as horizontal displacement velocities are comparable to velocity fluctuations characterizing high turbulence and thermal buoyancy.

Given the distance and time dispersion scales (< 3 meters and < 2-3 minutes) we can approximate the ECA as an airflow at constant atmospheric pressure, air density and dynamical viscosity ρ_*a*_ and *µ*. For a jet source (vaper’s mouth) approximated as an orifice of 1.5 − 3 cm^2^ area Gupta et al. (2010) (diameter *d*_0_ =1.25-1.75 cm) and initial velocities *U*_0_ given by (4), exhalation Reynolds numbers *Re* = (ρ/*µ*)*U*_0_*d*_0_ = 600 − 4400 are in the transition between laminar and turbulent, values well below the high Reynolds numbers expected near a jet source Pope (2001); Rajaratnam (1976), but we are mostly concerned with the jet evolution and displacement (penetration) along horizontal distances *𝓏* ≫ *d*_0_. Other parameters to consider are the injection time *t*_exh_ = 2 – 5 seconds and a temperature gradient from exhalation (initial) *T* = 30° − 35° C (mouth temperature) into an assumed *T* = 20° C for the surrounding air. For such values and scales the starting jet can be regarded as isothermal with thermal buoyancy becoming relevant only in the puff stage (see Ghaem-Maghami (2006); Ghaem-Maghami and Johari (2010)).

It is well known (see Pope (2001); Rajaratnam (1976); Morton et al. (1994); Shin et al. (2017)) that steady and unsteady jet/puff systems can be well approximated by analytic models that assume axial symmetry and a self similar profile for the average centerline and radial components of the velocity field in cylindrical coordinates 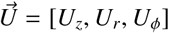 (see figure 1)

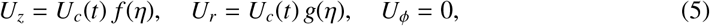

where *f, g* are empiric Gaussian or polynomial functions of the self similar variable η = *r*/*𝓏* and the centerline velocity is *U*_*c*_ = *U*_*𝓏*_ for *r* = 0 along the *z* axis, hence *f* (η), *g*(η) must satisfy *U*_*𝓏*_ = *U*_*c*_ and *U*_*r*_ = 0 at *r* = 0 (see examples in Wei and Li (2016); Pope (2001); Rajaratnam (1976); Abani and Reitz (2007); Abraham (1996); Ghaem-Maghami (2006); GhaemMaghami and Johari (2010); Sangras et al. (2002, 2003)). An axially symmetric self similar jet/puff system fulfills the conservation of linear specific momentum *Q* = *V U*_*c*_ (puff) and force 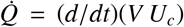 (jet) where *V* is the penetration volume Pope (2001); Sangras et al. (2002, 2003), hence 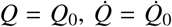 for an initial time *t* = *t*_0_. The stream wise centerline penetration distance and velocity for the jet and puff stages can be given by Sangras et al. (2002, 2003):

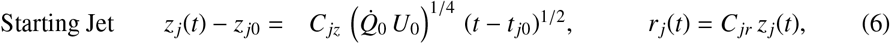

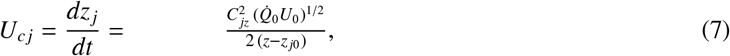

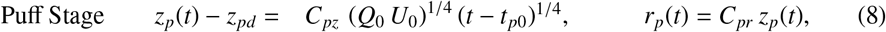

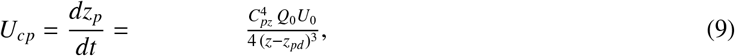

where the constants *C*_*j𝓏*_, *C*_*jr*_, *C*_*p𝓏*_, *C*_*pr*_ are empirically determined, and *𝓏*_*j*0_ is the *𝓏* coordinate value of the ejection orifice and *𝓏*_*pd*_ is the virtual origins of the puff (see Figure 2), which is an appropriate parameter to separate the starting jet and puff stages though it lies within the starting jet region (see detailed explanation in Sangras et al. (2002)). For the axial geometry of the jet/puff system under consideration we have 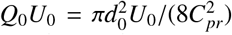 and 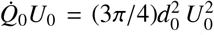. Following Sangras et al. (2002, 2003), we will choose the following numerical values for the constants in (6)–(9):

**Figure 2:**
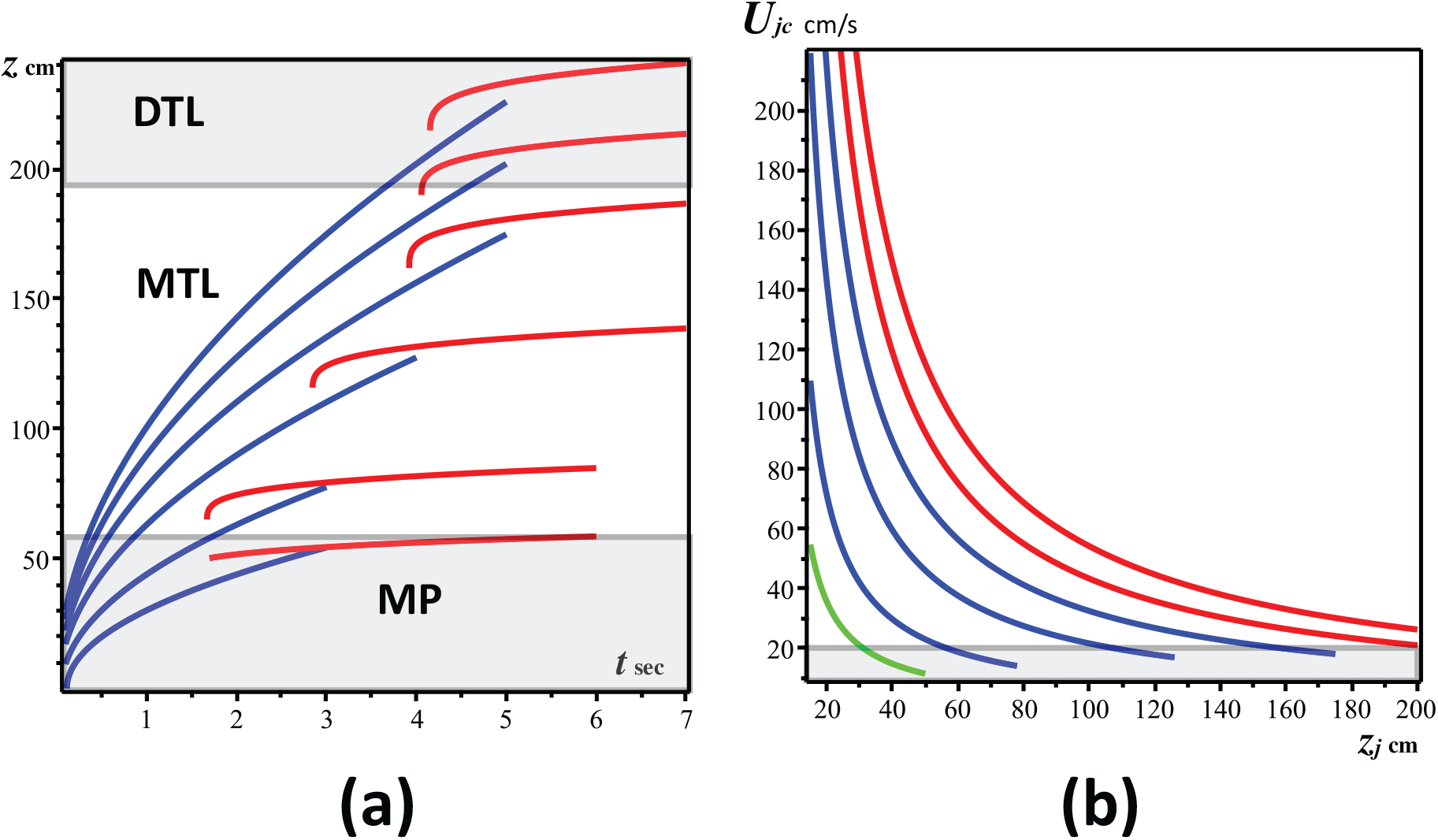
Jet/Puff horizontal displacement and centerline velocity. Panel (a) displays the displacement *𝓏* _*jc*_ of the staring jet (blue) and *𝓏*_*pc*_ of the puff (red) as functions of time from (equations (6)–(9)), for the three vaping topographies described in section 2.1: DTL (Direct to Lung), MTL (Mouth to Lung) and MP (Mouth Puffing). We assumed as injection (exhalation times) 3, 4 and 5 seconds. The initial velocities from bottom to top are *U*_0_ = 50, 100, 200, 300, 400, 500 cm/s. Panel (b) depicts centerline velocities *U*_*jc*_ for the starting jet (equation (7)), as functions of the horizontal displacement *𝓏*_*jc*_ during the injection times and initial velocities of panel (a) (green for MP, blue for MTL and red for DTL). Notice that once injection stops the jet has reached velocities ∼15− 20 cm/sec comparable to those of indoor air currents (bottom rectangle in (b)) and evolves into a puff.

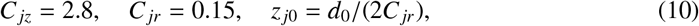

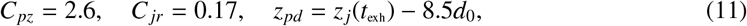

where the time *t*_*pd*_ and position of the puff virtual origin *𝓏*_*pd*_ is determined numerically from (6) by the condition *𝓏*_*j*_(*t*_*pd*_) = *𝓏*_*pd*_ (see Sangras et al. (2002, 2003)). Many vapers exhale at a downward angle typically γ ∼ 30 degrees, thus reducing the horizontal penetration of the starting jet given by (6) roughly as *𝓏*_*j*_ cos γ. From the dynamical equations (6)–(9) with the parameter values in (10)–(11) we display in section 5.2 the horizontal distance and jet/puff velocities characterizing direct exposure.

## 5. Results

### 5.1. Respiratory droplets emission

From the evidence and data examined in section 3 it is plausible to assume that droplet emission in smoking and vaping (at least MTL style) can be reasonably inferred from outcomes of studies in Table 2 with comparable exhaled tidal volumes (see Table SM(2), Supplemental Material), including outcomes of studies mentioned previously that examined breath holds.

The studies we have summarized and listed in Table 2 only involve mouth breathing, but share some common respiratory features with vaping and smoking: oral inspiration with usage of MP’s (in vaping), as well as qualitatively similar exhalation velocities and respiratory parameters: inhalation/exhalation times and tidal volumes. However, there are also differences: smoking and vaping do not involve the nose occlusion of these experiments, but involve suction which the subjects of the latter experiments did not experience. While absence of NC’s would imply a tidal volume very close to rest values in MTL smoking and vaping, this absence is compensated by the increase due to the need to overcome airflow resistance through suction. The decrease of droplet emission from the mouth/oropharynx hold in MTL topography (absent in normal breathing) was a detected outcome in two of the studies listed in Table 2. We have then the following inferences regarding emission of respiratory droplets

- **MTL vaping and smoking** (and even DTL vaping not involving deep inspiration). The outcomes displayed in Table SM(2) (Supplemental Material) and Table 2 suggest that exhaled droplets in mean tidal volumes *V*_*T*_ = 700 − 900 cm^3^, overwhelmingly in the submicron range (typically peaking at *d*_*p*_ = 0.3 − 0.8 *µ*m), as well as a small rate of droplet emission: roughly *N*_*p*_ = 6 − 200 per exhalation (mean *N*_*p*_ = 79.82, standard deviation 74.66, computed from the outcomes listed in Table 2 for exhaled volume of 800 cm^3^), with droplet number densities well below *n*_*p*_ = 1 cm^−3^. However, the wide individual variation reported in these studies should also apply to vaping, including the existence of a small minority of outlier individuals that can be thought of as “super emitters” reaching over *N*_*p*_ ∼ 1000 per exhalation.
- **DTL vaping**. It involves a spectrum of deeper respiratory intensity than MTL vaping and thus should involve a higher rate of droplet emission. Perhaps the closest analogue in the studies listed in Table 2 to infer droplet emission for intense DTL (2-3 LT exhalation) breathing at fractional residual capacity in Almstrand et al. (2010) that reported emission rates of around 1000/LT. However, this style of vaping is practiced by a small minority of vapers (roughly 10-20%, see figure SM(1), Supplemental Material), while extreme vaping with big clouds (the so called “cloud chasers”) is even less frequently practiced in competitions or exhibitions. Evidently, this type of extreme vaping cannot be sustained for long periods and is not representative even of DTL vapers.

While the inferred droplet numbers in the upper end of high intensity DTL vaping can be comparable with low end numbers for vocalizing, the latter involves modes with larger mean diameters because of distinct droplet generation processes (see Asadi et al. (2019); Morawska et al. (2009); Johnson et al. (2011)).

### 5.2. Distance for direct exposure

Direct exposure to respiratory droplets carried by exhaled ECA can be inferred from the horizontal displacement or penetration distance of the jet/puff system whose dynamics follows from equations (6)–(9) with the parameter values in (10)–(11). We display in figure 2 displacement distances and centerline velocities for assorted values of initial exhalation velocities *U*_0_ corresponding to the vaping intensities we have considered.

Notice that the maximal penetration goes beyond that afforded by the momentum trust of the starting jet, with the puff further evolving at lesser speeds. Horizontal penetration varies from 0.5 meters for Mouth Puffing (*U*_0_ = 0.5 m/s) through the range between 0.6 and 2.0 meters the MTL regime (*U*_0_ = 0.5 − 3.75 m/s) and beyond 2 meters for the higher intensity DTL regime (*U*_0_ = 1.5 − 5 m/s). Centerline velocity drops to about 0.2 m/s at different times and distances when fluid injection stops in all cases.

Given its short time duration and close distance scope of the momentum trusted staring jet, the analytic model (6)–(7) remains a reasonably good approximation to infer the necessary distance to minimize the risk of direct exposure of bystanders to respiratory droplets. As the jet evolves while fluid is injected there is increasing entrainment from the surrounding air at velocity *U*_*e*_ ∝ *U*_*r*_, with entrained air reaching about 40% of the jet mass at the end of injection in the transition towards the puff (around its virtual origin, see Ghaem-Maghami (2006); GhaemMaghami and Johari (2010)). Since there are airflow currents of ∼ 10 cm/s (and up to 25 cm/s) even in still air in home environments with natural ventilation Matthews et al. (1989); Berlanga et al. (2017), at this stage the puff formation can be easily destabilized by vortex motion generated through turbulent mixing from the large velocity fluctuations produced by the entrainment (see details in Wei and Li (2015); Vuorinen et al. (2020)).

Turbulence and thermal buoyancy become important factors when there is human motion or walking Wang and Chow (2011), or in micro-environments with mechanical ventilation (mixed or displaced), resulting in a faster disruption and dispersion of the slow moving puff, carrying the submicron ECA and respiratory droplets along the air flow. In general, submicron droplets exhaled at the velocities under consideration can remain buoyant for several hours, with mixing ventilation tending to uniformly spread them, whereas directed ventilation tends to stratify them along different temperature layers (see comprehensive treatment by He et al. (2011); Gao and Niu (2007); Gao et al. (2008)). In all cases there is a risk of indirect contagion by exposure to these droplets. The detailed description of droplet dispersion after the puff is disrupted is a complicated process that requires computational techniques that are beyond the scope of this paper (see comprehensive analysis by Vuorinen et al. (2020)).

## 6. Limitations, final discussion and conclusion

We have presented in this paper a comprehensive analysis and theoretical modeling of the plausibility, scope and risk for pathogen (including SARS-CoV-2 virus) contagion through direct and indirect exposure to respiratory droplets that would be carried by ECA (e-cigarette aerosol) exhaled by vapers. An summary that outlines the methodological structure and obtained results of the article is provided in the Introduction.

### 6.1. Limitations

#### 6.1.1. Lack of empiric data

It is important to openly recognize the main limitation of this study: the lack of experimental and observational data on respiratory droplets carried by exhaled ECA. It is quite plausible that emission of these droplets should occur, as exhaled ECA is an expiratory activity, but without empiric data any quantitative assessment of its nature and scope must necessarily be inferred or estimated indirectly, either through theoretical speculation from the physical and chemical properties of ECA, or through extrapolation from available data on other expiratory activities that can serve as reasonable proxies for vaping. The need to provide the best possible and self consistent inference on this missing data explains and justifies the length of the present study: data availability would render several sections (for example sections 3.1, 3.2 and 3.3) redundant or drastically shortened and kept only for comparative reference.

#### 6.1.2. Oversimplification of vaping styles

The classification of puffing topographies in two separate mutually exclusive categories (MTL and DTL) that we presented in section 2.1 roughly conveys the two main vaping styles, but ecigarettes are a rapidly changing technology and thus this simplified approach cannot capture the full range and scope of individual vaping habits.

#### 6.1.3. Oversimplification of infective parameters and individual variability

We remark that the ranges of numerical values we have obtained of emitted droplets possibly emitted by vaping are rough average estimates gathered from outcomes reported in breathing studies (listed in Table 2) involving a wide variety of subjects, including both healthy and individuals affected by respiratory conditions (not by SARS-CoV-2). We have not considered the small minority of outlier individuals who are super spreaders emitting significantly larger numbers of droplets Asadi et al. (2019). We have also considered simply droplet emission, disregarding the specification of a specific pathogen. Evidently, this oversimplification disregards important known facts, for example: droplet characteristics vary among pathogens and between healthy and infected subjects Fabian et al. (2011); Wurie et al. (2013).

In fact, numerous aspects associated with the spreading and infection details of the SARS-CoV-2 virus remain uncertain and subject to large (often unexplained) individual and environmental variability (a good summary of these uncertainties is found in Klompas et al. (2020); Morawska and Milton (2020); Morawska and Cao (2020); NAS (2020)). However, in order to be able to model a possible (previously unexplored) route of droplet transmission and possible infection, it is necessary and unavoidable to simplify this complexity and lack of data to obtain plausible order of magnitude estimates that can be verified once empiric evidence is available.

#### 6.1.4. Oversimplification of droplet dynamics

Since respiratory carried by exhaled ECA are expected to be overwhelmingly submicron, and thus carried by the fluid flow, the simple dynamical modeling of a starting jet followed by an unstable puff (section 4) is sufficient to estimate direct exposure distances. However, we recognize its limitations: it is strictly valid for a jet/puff system emitted by a static vaper in typical indoor conditions with idealized natural ventilation. Evidently, to estimate the fluid flows that determine this exposure (and indirect exposure by dispersing droplets) in less idealized conditions requires a more realistic description using computational methods of fluid mechanics to incorporate effects of turbulence and thermal bouyance, as well as air currents from ventilation or motion. Rather, we examine global volume exposure in a separate article through a risk model not involving fluid dynamics described in Sussman et al. (2020). It is important to mention that this simplification of the dynamics is harder to justify for expiratory activities like coughing or sneezing, as the latter involve larger ejection velocities and a much wider spectrum of droplet diameters that includes significant number of large supermicron droplets (significant numbers of diameters 1−10 *µ*m and even > 100 *µ*m) whose effect on the dynamics of the carrier fluid cannot be neglected (these are strictly speaking multiphasic flows Yeoh and Tu (2019); Scharfman et al. (2016); Bourouiba et al. (2014)).

### 6.2. Safety considerations

#### 6.2.1. Respiratory flow visualization

As opposed to other respiratory activities (speaking, singing, coughing, sneezing), the involved respiratory flow of ECA is visible because the carried submicron droplets (ECA and respiratory) act effectively as visual tracers of the carrier fluid (see sections 2.3 and further detail in Sussman et al. (2020)). Besides the evident psychological dimension of this flow visualization, there are safety implications: vapers and those surrounding them have a clear, instinctive and immediate delineation of the flow’s horizontal distance reach and spreading direction along the exhaled jet. From the outcomes of our hydrodynamical analysis (section 5.2), we can recommend as a basic safety measure to avoid direct exposure (irrespective of face mask wearing) by keeping a 2 meter distance away from the vaper (when vaping) in the direction of the visible jet. In other directions the exposure would be indirect, but nevertheless it is prudent to maintain 2 meters of separation in all directions from anyone vaping when not wearing a face mask. Notice that these recommended safety measures coincide with the standard social separation recommendations adopted worldwide Hsiang et al. (2020).

#### 6.2.2. Final conclusion

This article is the first comprehensive attempt (as far as we are aware) to assess the plausibility and distance range of possible transport by exhaled ECA of respiratory droplets potentially carrying pathogens (including the SARS-CoV-2 virus). Given the lack of empiric data on this phenomenon we utilized data from respiratory parameters of cigarette smoking and droplet emission from mouth breathing, both considered as valid proxies for vaping exhalations. Our results can provide useful guidelines to address the possible scope of vaping exhalations in assessing risks of contagion of infectious disease in shared indoor spaces. In other articles we have examined actual contagion risks and public policy implications (see Sussman et al. (2020, 2021)). Setting aside harms from environmental tobacco smoke unrelated to COVID-19, these results also apply to sharing an indoor space with a smoker.

## Data Availability

The data that support the findings of this study are available from the corresponding author, RAS, upon reasonable request.

## Footnotes

1 The 5 *µ*m cut–off separating larger droplets and “aerosols” is merely a convention that artificially simplifies droplet dynamics that vary along a continuous spectrum of diameters into two mutually exclusive modalities. We will avoid altogether the “droplets” vs “aerosols” terminology, with the term “droplets” referring henceforth to generic respiratory droplets of continuously varying diameters.

2 A search of the literature revealed three opinion pieces: Ahmed et al. (2020); Sifat et al. (2020); Mahabee-Gittens et al. (2020) that offer a very limited argumentation.

3 This paper will not address potential COVID-19 contagion through respiratory droplets carried by environmental tobacco smoke, though smoking can serve as a useful proxy for understanding the respiratory and dynamical parameters of low intensity (‘mouth to lung’) puffing style practiced by 80-90% of vapers. However, most of the results we obtain are applicable to “mainstream” smoke exhalations emitted by smokers, not to sidestream emissions from the burning/smouldering tip of cigarettes, cigars and pipes that make the bulk of environmental tobacco smoke.

4 These machine puff time lapses are different from those reported in Table 1. The former correspond only to inhalation times as instruments aim at simulation of a mouth inhalation, the latter are time lapses in human vapers and thus include inhalation and exhalation.

