## Supplemental Material (1) for "Modeling aerial transmission of pathogens (including the SARS-CoV-2 virus) through aerosol emissions from e-cigarettes"

**Supplemental Material (1) Usage of different classes of e-cigarette devices in the US and UK markets**

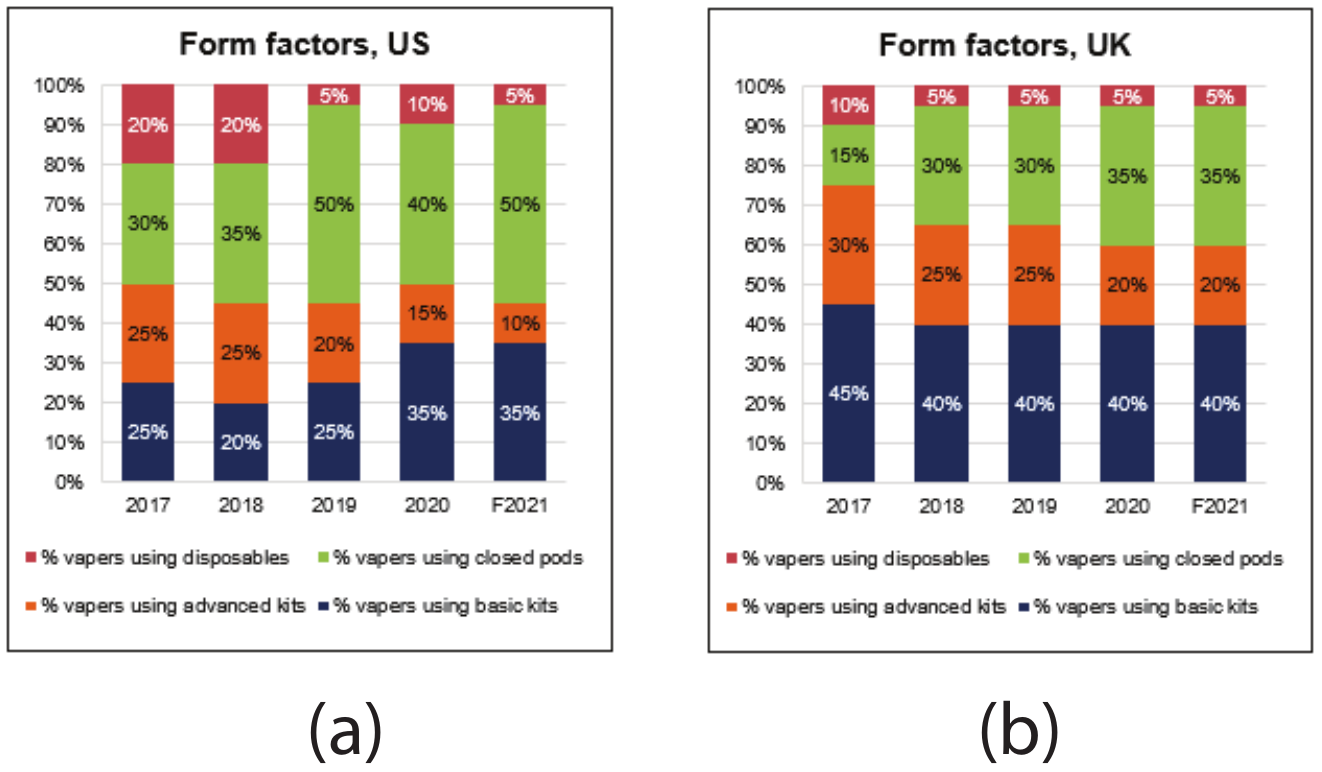

**Figure SM(1) 1**. Disposables, closed pods and basic kits are all low intensity vaping devices appropriate for the low intensity Mouth to Lung (MTL) puffing topography (vaping style) similar to cigarette smoking. Notice that in 2020 only 15 % and 20% of consumers in the USA and the UK used advanced kits that allow for the intense Direct to Lung (DTL) vaping style. Credit to Ecig intelligence: Databases - key global analysis of the vapour sector. Https://ecigintelligence.com/content types/database/, retrieved October 28, 2020

The USA and the UK are the largest and most developed e-cigarette markets, a fact that explains why the closed system category is more prevalent. In a natural evolution of markets, the vapor category takes off with a more hobbyist segment of users who are more likely to vape with Direct to Lung (DTL) topography in high powered devices that yield large clouds. In nascent markets the “easy to use” open system devices are not of great quality, though recent innovations are likely to improve this. Smokers in large markets are also likely to have higher disposable incomes and a more developed attitude of willing to (and being able to afford to) switch to a less harmful alternative. Such markets also have extensive distribution networks (convenience stores, tobacconists, etc.). These factors influence the dominance of the market share of closed system devices and thus to characterize low intensity Mouth to Lung (MTL) style similar to cigarette smoking as the most prevalent among the vast majority of vapers.
