## Supplemental Material (2) for "Modeling aerial transmission of pathogens (including the SARS-CoV-2 virus) through aerosol emissions from e-cigarettes"

**Supplemental Material (2)**. Respiratory parameters of cigarette smoking

| **Study authors & reference** | **Inhalation/Exhalation Volume ($T_V$ in mL)** | **Puff time/volume** PT seconds, PV mL | **Comments** |
|---|---|---|---|
| Bernstein (2004) | 591 (mean), 560 (median)<br>Range $413-918$ | PT 1.8 (mean)<br>PT Range 1.6-2.4<br>PV 43 (mean)<br>PV Range 21-66 | Summary of 32 studies before 1988<br>US Surgeon Report 1988 |
| Tobin et al. (1982b) | $841 \pm 517^*$ Natural<br>$748 \pm 323^{**}$ Natural<br>$878 \pm 431^*$ Cig Holder<br>$815 \pm 376^{**}$ Cig Holder<br>Range 270-1990 mL | | 10 subjects<br>Non invasive RIP |
| Tobin et al. (1982a) | $790 \pm 450$ Group Average<br>$460 \pm 130$ Rest Tidal Vol<br>Range 270-1970 mL | PT $4.5 \pm 1.3$<br>Includes Breath Hold | 19 subjects<br>Non invasive RIP |
| Nil et al. (1986) | $500 \pm 300^*$ Men<br>$600 \pm 500^{**}$ Men<br>$400 \pm 300^*$ Women<br>$400 \pm 300^{**}$ Women | PV $42.3 \pm 14.5$<br>PV $50.2 \pm 16.8$<br>PV $41.4 \pm 13.3$<br>PV $47.0 \pm 15.8$ | 67 men, 48 women |
| Woodman et al. (1986) | 192-644 Total Inh Smoke<br>315-919 Total Inh Vol | PT 1.2-2.9 | Inert Krypton gas as smoke tracer |
| Robinson et al. (1992) | $828 \pm 126$ Low Nicotine<br>$845 \pm 105$ Normal Nicotine | | |
| Charles et al. (2009) | $833 \pm 279$ Inhaled Vol<br>$897 \pm 308$ Exhaled Vol<br>$500 \pm 148$ Rest Tidal Vol | $1.82 \pm 1.16$ Inh Time<br>$2.28 \pm 0.87$ Exh Time | 74 subjects<br>Non invasive RIP<br>without Cig Holder |
| Marian et al. (2009) | $702 \pm 437^*$ Inh Vol<br>$636 \pm 138^{**}$ Inh Vol<br>$577 \pm 329^*$ Exh Vol<br>$655 \pm 195^{**}$ Exh Vol | $1.19 \pm 0.29^*$, Inh Time<br>$1.22 \pm 0.37^{**}$, Inh Time<br>$2.01 \pm 0.76^*$, Exh Time<br>$2.89 \pm 0.72^*$, Exh Time<br>$0.45 \pm 0.48^*$, Breath Hold<br>$0.45 \pm 0.57^{**}$, Breath Hold<br>PV $44.9 \pm 12.3^*$<br>PV $44.5 \pm 10.9^{**}$ | BAT study 1986<br>Table 2 |

**Table SM(2)**. The table lists various inhaled/exhaled volumes and associated puff times and volumes. The term “puff time” (PT) denotes the time taken to draw smoke from the cigarette (puffing) with “puff volume” (PV) denoting the drawn volume before it mixes with air. Volumes in the second column refer to the inhaled mixture of smoke and air unless it is explicitly specified that it refers to the exhaled mixture. The symbols ±, * and ** respectively denote standard deviation, high and low TAR yields. RIP refers to Respiratory Inductive Plethysmograph, BAT is British American Tobacco.

**Supplemental Material (2). Respiratory parameters of cigarette smoking**
